# AI-powered Gradient Echo Plural Contrast Imaging (AI-GEPCI) – a Comprehensive Multiparametric Neurological Protocol from a Single MRI Scan

**DOI:** 10.64898/2026.02.11.26346017

**Authors:** Jeramy Lewis, Manu S. Goyal, Gregory F. Wu, Yuyang Hu, Alexander L. Sukstanskii, Satya V.V.N. Kothapalli, Anne H. Cross, Ulugbek Kamilov, Dmitriy A. Yablonskiy

**Affiliations:** Mallinckrodt Institute of Radiology, Washington University in St. Louis; Department of Neurology, Washington University in St. Louis; Department of Electrical & Systems Engineering, Washington University in St. Louis; Department of Computer Science & Engineering, Washington University in St. Louis, Washington University in St. Louis School of Medicine, 660 S. Euclid Ave, St. Louis, MO, USA

**Keywords:** Gradient Echo Plural Contrast Imaging (GEPCI), Magnetic Resonance Imaging (MRI), Attention Convolutional Neural Networks (ACNN), Central Vein Sign (CVS), Paramagnetic Rim Lesions (PRL), Multiple Sclerosis (MS)

## Abstract

**Background:** MRI plays an essential role in diagnosing and monitoring neurological diseases. Conventional protocols rely on multiple sequences to obtain complementary contrasts, increasing scan time, cost, and tolerability. Generating multiple contrasts from a single acquisition may streamline workflow while maintaining clinical utility.

**Purpose:** Train attention-based convolutional neural networks (ACNNs) to generate clinical-quality FLAIR, MPRAGE, R2*, and derived contrasts from a single Gradient Echo Plural Contrast Imaging (GEPCI) acquisition, enabling multi-contrast imaging from one scan.

**Study Type:** Retrospective.

**Population:** 43 MRI scans from individuals with multiple sclerosis (25/18 F/M, 49±11 years old).

**Field Strength/Sequence:** 3T MRI was used to obtain 3D GEPCI, MPRAGE, and FLAIR sequences.

**Assessment:** Technical quality of the AI-generated contrasts was evaluated against directly acquired MRI images using structural similarity index (SSIM). Quantitative accuracy for R2* maps was evaluated using normalized root-mean-square error (NRMSE). Clinical image quality was assessed by expert physicians. Lesion volumes and counts were obtained using automated segmentation.

**Results:** AI-generated FLAIR and MPRAGE images achieved mean SSIM values of 0.923±0.028 and 0.935±0.022, respectively. The generated R2* maps achieved a mean SSIM of 0.996±0.006, with quantitative accuracy reflected by an NRMSE of 0.031±0.020. Physicians rated GEPCI-FLAIR images at 4.2 and GEPCI-MPRAGE images at 4.5 (on a 1-to-5 scale), both exceeding the clinically routine standard of 4.0. Lesion volume and count comparisons from automated segmentation showed strong agreement between AI-generated and ground-truth measurements (R²=0.988 and R²=0.933, respectively).

**Conclusion:** AI-GEPCI generated multiple clinically relevant MRI contrasts from a single GEPCI acquisition with high similarity to corresponding acquired images. Radiological reviews and quantitative analyses supported the feasibility of producing high-quality, intrinsically co-registered multi-contrasts for comprehensive brain evaluation.

## Introduction

Machine learning (ML) methods have increasingly been applied to medical image reconstruction and analysis, providing data-driven approaches for generating or enhancing MRI contrasts. A typical MRI examination consists of multiple pulse sequences, each providing a distinct image contrast sensitive to different tissue properties. Alternative methods use MRI pulse sequences capable of generating multiple contrasts from a single acquisition, such as MR fingerprinting (1) and Gradient Echo Plural Contrast Imaging (GEPCI) (2). The improved image quality and reduced computation times associated with these approaches can be achieved through ML based reconstruction techniques (3, 4). The present work is centered on further developing the GEPCI technique by introducing artificial intelligence powered GEPCI (AI-GEPCI), a deep learning extension of the original GEPCI method (2), aimed at accelerating image reconstruction while enhancing the rich contrast and quantitative information GEPCI offers.

GEPCI is a post-processing technique that extracts several image contrasts from a single Gradient Recalled Echo (GRE) MRI scan with multiple gradient echo (mGRE) pulse sequences (2) readily available on most commercial MRI scanners. It was previously demonstrated that GEPCI reconstruction algorithms (2) allow the generation of images like T1-weighted, T2-weighted FLAIR (Fluid-Attenuated Inversion Recovery), and SWI (Susceptibility Weighted Imaging), along with quantitative R2* (1/T2*) maps, phase, and QSM (Quantitative Susceptibility Mapping) images. An advanced version of GEPCI, quantitative Gradient Recalled Echo (qGRE) (5, 6), allows further separation of the R2* relaxation rate parameter into its subcomponents: R2t* - sensitive to tissue microstructure, and R2’ - sensitive to vascular BOLD (Blood Oxygen Level Dependent) signal properties, both corrected for macroscopic field inhomogeneities (7) and physiological fluctuations (8). GEPCI and qGRE have proved useful in studying neurodegenerative diseases, such as Alzheimer’s Disease (9, 10), multiple sclerosis (MS) (8, 11–14), psychiatric diseases (15), and traumatic brain injury (16). While the classical GEPCI pipeline employs a multi-step process with model-based fitting and investigator-guided reconstructions, the introduced herein AI-GEPCI approach leverages deep learning to automatically extract key features from the GEPCI images and perform reconstructions orders of magnitude faster, with enhanced lesion visualization and superior quantitative accuracy.

This study aimed to demonstrate that attention-based convolutional neural networks (ACNNs) (17) can generate clinical-quality FLAIR, MPRAGE, and R2* images along with their derived contrasts, including T2*-weighted images, FLAIR* (18), and introduced herein MP*, from a single GEPCI acquisition. Individuals with MS, a chronic inflammatory disease of the central nervous system characterized by demyelination, axonal injury, and neurodegeneration (19–22), routinely undergo MRI for diagnosis and monitoring using sequences such as FLAIR, MPRAGE, SWI, and recently added FLAIR*, which together capture lesion visibility, structural detail, and overall lesion burden (23, 24). This study demonstrates that GEPCI magnitude data could serve as the sole input for ACNN-based synthesis of these clinically relevant contrasts, enabling a rapid, intrinsically co-registered, and multiparametric imaging workflow suitable for comprehensive MS lesion evaluation, including biomarkers such as the central vein sign (CVS) and paramagnetic rim lesion (PRL) features, within a single mGRE scan (25–31).

## Materials and Methods

This retrospective cohort study, approved by the Washington University IRB, consisted of 43 MRI scans from individuals with a clinical diagnosis of MS (25/18 F/M, 49 ±11 years old) who had concurrently acquired GEPCI, FLAIR, and MPRAGE sequences available for analysis. No inclusion or exclusion criteria were applied beyond the existence of complete GEPCI, FLAIR, and MPRAGE datasets. The 43 available scans were randomly partitioned into training (n = 24), validation (n = 3), and testing (n = 16) subsets.

### Imaging Protocol and Ground Truth Data Generation

MRI data were acquired with a Siemens-3T Prisma scanner (Siemens Healthineers, Erlangen, Germany) equipped with a 32-channel Radio Frequency (RF) coil. FLAIR and MPRAGE used standard Siemens sequences with FOV of 256 mm, resolution of 1×1×1 mm^3^ and GRAPPA (32) algorithm with acceleration factors of 2 in phase and slab directions. MPRAGE was acquired with TR/TE/TI = 2300/2.98/900 msec. Parameters for FLAIR were TR/TE/TI = 5000/393/1800 msec. GEPCI sequence had the same FOV of 256 mm, resolution of 1×1×1 mm^3^ and GRAPPA algorithm with acceleration factors of 2 in phase and slab directions. Other GEPCI parameters were: TR=50 msec, 10 gradient echoes with first TE=4 msec and echo spacing = 4 msec, FA=30.

After correction for physiological fluctuations (8) and GRAPPA reconstruction (32), the 32-channel data were combined using a previously developed algorithm (2). The quantitative R2* maps were produced by voxel-by-voxel fitting the biophysical model to the 10 GRE images:

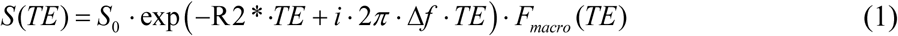

In (Eq. 1), *TE* is the gradient echo time, *S*_0_ is the signal amplitude, Δ*f* is the local frequency shift, *F_macro_(TE)* accounts for the adverse effects of macroscopic magnetic field inhomogeneities by means of a voxel spread function method (7).

The FLAIR and MPRAGE images were co-registered to the GEPCI using the FMRIB Software Library (FSL) (33). The magnitude of the complex GEPCI data was used and normalized by dividing all echoes by the mean value calculated across all the first echo images. The FLAIR and MPRAGE volumes were normalized to the mean value of their respective datasets.

### Network Architecture and Training

The ACNNs utilized the attention U-Net architecture (17) consisting of a down-sampling encoder path and an up-sampling decoder path incorporating attention blocks (Figs. 1, 2). The 3D volumes are loaded as sets of transverse 2D slices. Once loaded, the encoder processes all slices (10 echoes per slice) of the 3D GEPCI volume simultaneously, extracting multi-scale features. The decoder up-samples the feature maps and refines the images utilizing the attention blocks and skip connections to combine information from corresponding encoder layers. The final convolutional layer produces a single reconstructed output 3D image volume. By integrating attention blocks within the decoder, the network learns to selectively focus on echoes based on their relaxation-weighted information, emphasizing earlier echoes for their T1-weighted features (beneficial for MPRAGE translation) and later echoes for their T2-weighted contrast (important for FLAIR reconstruction). The attention gates dynamically modulate feature responses by emphasizing salient spatial and echo-dependent patterns while suppressing less informative signals. In medical imaging, this selective focus has been shown to improve network interpretability and performance in tasks such as segmentation, classification, and modality translation (34–36). Given the echo-dependent tissue contrast variation and lesion heterogeneity inherent in GEPCI, the attention blocks help the network prioritize features from echoes that contribute most to accurate synthesis.

**Figure 1.**
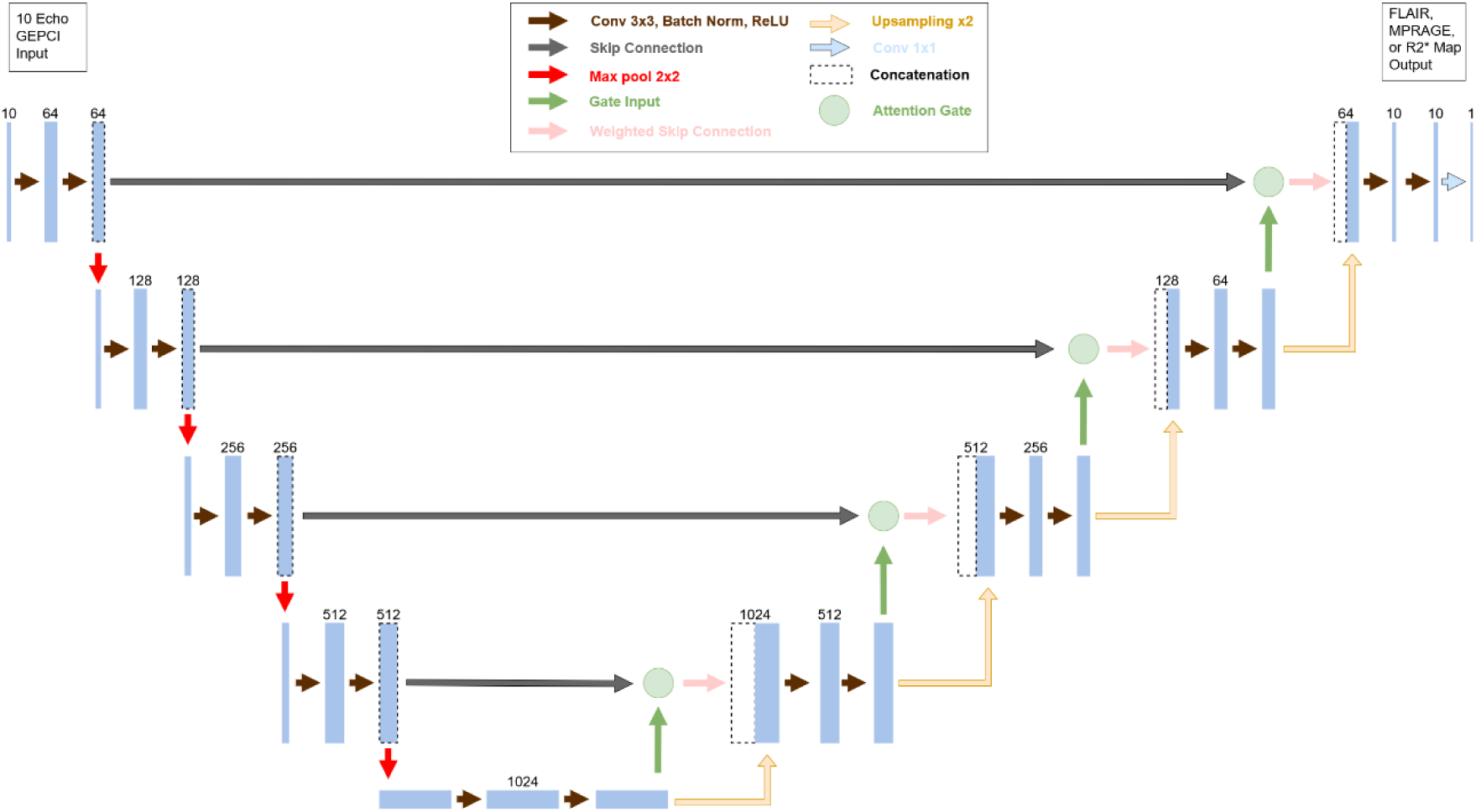
Network diagram for the attention U-Net architectures utilized in this study (17). The FLAIR, MPRAGE, and R2* map networks use the same architecture with different parameters. The networks take a 2D slice from a 3D GEPCI MRI volume with 10 gradient echoes as input. Each encoding block for the downsampling path performs two 3×3 convolutions, batch normalization, ReLu activation, and a 2×2 max pooling operation. The feature maps from each encoder block are passed forward via skip connections to the corresponding decoder block in the upsampling path. In the upsampling path, the decoder blocks utilize attention gates (detailed in Fig. 2). The weighted skip connection from the attention gate and the two times upsampled output from the previous block are concatenated, have two 3×3 convolutions, batch normalization, and ReLu activation. The final decoder block includes an additional 1×1 convolution to reduce the 10 input channels to a single output image.

**Figure 2.**
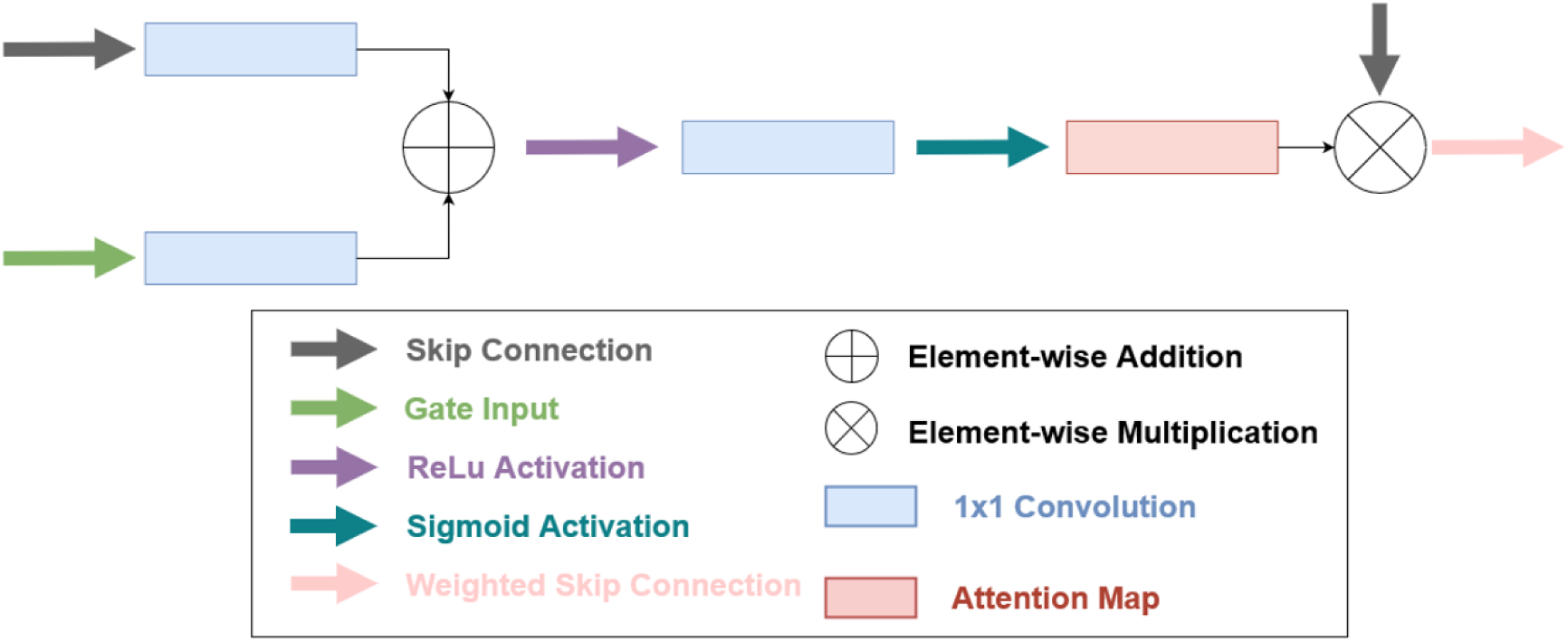
Detailed attention gate architecture. The attention gates receive the upsampled output from the previous decoder as the gating signal and the feature map from the corresponding encoder skip connection. Each map is processed individually through a 1×1 convolution, added elementwise, and activated by ReLu. The result is passed through another 1×1 convolution followed by a sigmoid function to produce a spatial attention map. This attention map is multiplied elementwise with the original skip connection, allowing only the most relevant features to pass to the decoder block via concatenation.

During upsampling decoding steps, the attention gates weight features from different echoes according to their relevance, enabling the network to combine multi-scale representations from the encoder in a structured manner.

Two input configurations were evaluated for network training. In the primary configuration, whole-head GEPCI magnitude volumes were used as inputs, and the corresponding whole-head FLAIR, MPRAGE, or R2* images served as target ground truths. In a secondary configuration, brain-extracted versions of these volumes were used, in which non-brain voxels were set to zero based on the brain-extraction mask, and the corresponding brain-extracted target images were used as labels. Unless otherwise specified, the results reported in this work refer to models trained using whole-head inputs and labels.

To assess the effect of attention mechanisms, a post-hoc ablation experiment was performed in which the ACNN architecture was compared with an otherwise identical CNN model lacking attention gates. Both models were trained under the same conditions, using identical training, validation, and testing splits, loss functions, learning rates, and preprocessing steps. Performance was evaluated using Structural Similarity Index Measure (SSIM) (37) for the FLAIR, MPRAGE, and R2* map outputs and normalized mean absolute error (NMAE) for the R2* maps as well.

The models were developed in Python-3.10.13 using PyTorch-2.0.0 and trained on a Windows-10 Enterprise machine equipped with an NVIDIA RTX A6000 GPU (compute capability of 8.6). Training was conducted for 1,000 epochs with a batch size of 2 using the Adam optimizer. The following loss functions were evaluated: L2 (mean squared error), L1 (mean absolute error), SSIM, and a combination of 50% weighted L2 with 50% weighted SSIM. For the ground truth, the networks were trained with and without using the FSL brain extraction tool (BET). Experiments with various learning rates were performed to select the combination yielding the best training and validation losses. For GEPCI-to-FLAIR and GEPCI-to-MPRAGE networks, the learning rate was set at 10^-5^, while for GEPCI-to-R2* it was set at 4 ·10^−4^. The attention blocks employed typical parameters: kernel size 1×1, stride 1, and no padding.

### GEPCI-derived Images

Derived contrasts were generated from the ACNN-produced battery of GEPCI images. By using quantitative R2* maps, T2*-weighted images (T2*WI) (Eq. 2), FLAIR*, and MP* images (Eq. 3) can be computed for arbitrary echo time TE:

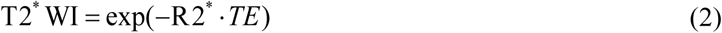

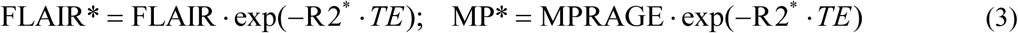

These TE-dependent images were used to evaluate how echo time influences contrast behavior. Unless otherwise specified, example derived images in the Results section correspond to TE = 50 msec.

### Assessment of Central Vein Sign (CVS) and Paramagnetic Rim Lesions (PRL)

CVS and PRL assessments were performed using qualitative visual evaluation based on established criteria for MS lesion characterization. A lesion was considered to demonstrate a CVS if a linear structure consistent with a small vein appeared hyperintense on FLAIR* and was visible in at least two imaging planes, following the guidelines in (30). CVS also appeared hyperintense on T2*WI, FLAIR*, and MP*, and hypointense on R2*. Similarly, a lesion was classified as a PRL if it exhibited a peripheral rim that appeared hypointense on FLAIR* in at least two orthogonal planes, consistent with accepted definitions (30). PRL was also characterized as peripheral rim that appeared hypointense on T2*WI, and MP*, and hyperintense on R2*. CVS and PRL evaluations were performed using the ACNN-generated R2* maps and the derived T2*WI, FLAIR*, and MP* images.

### MS Lesion Evaluation

Lesion masks were generated using the automated Lesion Segmentation Tool (LST) for Statistical Parametric Mapping (SPM) (www.statistical-modelling.de/lst.html), version 3.0.0. For most subjects, lesions were segmented using the lesion growth algorithm (LGA) (38) which combines T1-weighted tissue classification with FLAIR intensities to derive lesion probability maps. For two subjects, severe motion artifacts in the MPRAGE images were identified during this review process, and these cases were subsequently processed using the automated lesion prediction algorithm (LPA) (39) which uses a logistic regression model trained on FLAIR data and spatial lesion priors.

### Image Analysis

To assess the technical quality of the AI-generated MRI images, image similarity was evaluated only within brain tissue by masking out non-brain regions such as skull, scalp, and background. SSIM was computed between the generated FLAIR, MPRAGE, and R2* images (G-FLAIR, G-MPRAGE, and G-R2*) and their respective ground-truth images. The accuracy of the quantitative G-R2* maps was additionally evaluated using the normalized root-mean-square error (NRMSE), calculated relative to the mean R2* value across the brain. For data analysis, 3D images were reconstructed in all three planes (axial, sagittal, and coronal). All metrics were computed slice-by-slice within each orientation and then aggregated to yield a single whole-volume metric per subject. The results are reported as mean ± standard deviation for each test subject and across test subjects. While NRMSE was used for primary quantitative evaluation of R2* accuracy, NMAE was additionally employed in the ablation analysis to provide a complementary assessment of absolute error that is less sensitive to error squaring.

To evaluate lesion segmentation performance in the context of image-quality assessment, lesion volumes and lesion counts were obtained using fully automated SPM-based segmentation (LGA/LPA algorithms), and measurements from the AI-GEPCI generated images were compared with those from the corresponding ground-truth segmentations using scatter plots. Agreement between lesion volume and lesion count measurements obtained from the AI-generated images and the corresponding ground-truth segmentations was assessed using the coefficient of determination (R²). The lesion statistics included in this study were provided solely for descriptive comparison to support image-quality evaluation and were not intended as validation of a lesion-segmentation method, which was beyond the scope of the present work.

To assess the radiological quality of the generated brain MRI images, the brain-extracted G-FLAIR, G-MPRAGE, and corresponding ground-truth FLAIR and MPRAGE volumes were reviewed by three physicians: one neuroradiologist with 10 years of experience (M.G.) and two MS neurologists with 40 and 20 years of experience, respectively (A.C. and G.W.). Each subject’s GEPCI-generated and corresponding ground truth image volumes were labeled as either A and B or B and A in pseudo-randomized fashion and physicians independently reviewed each pair of blinded images according to overall quality, posterior fossa quality, and artifact quality on a 1 to 5 Likert scale, with 4 being equivalent to clinically routine imaging and 5 exceeding this quality. Physicians were asked to identify the GEPCI-generated images, and which (if either) was preferred. They also specified whether they could detect additional lesions on either the ground truth or GEPCI-generated images.

## Results

Three separate ACNNs were trained to generate G-FLAIR, G-MPRAGE, and G-R2* maps from original GEPCI data. The general performance results with different loss functions used for network training are summarized in Table 1. Among the evaluated loss functions, the L2 loss resulted in higher mean SSIM values and lower variability for the G-FLAIR and G-MPRAGE reconstructions compared with the other losses, based on descriptive metrics. The SSIM loss produced lower mean SSIM values and greater variability across test subjects. For G-MPRAGE, the L1 and L2 loss functions produced similar descriptive SSIM measures. For the quantitative R2* maps, only L1 and L2 loss functions were evaluated, and their descriptive performance metrics were comparable.

**Table 1.**
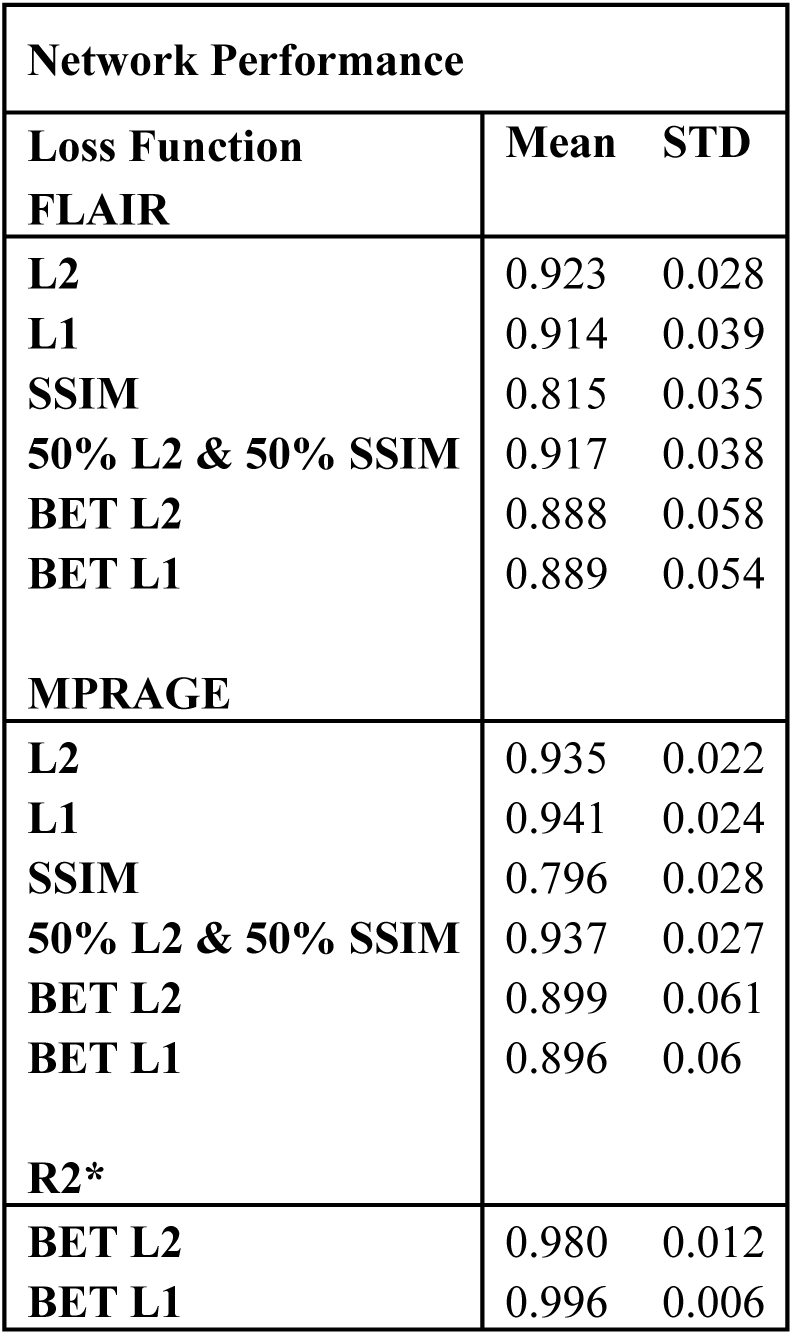
Results obtained with different loss functions. Data shows SSIM metric assessments (mean ± standard deviation across slices covering the brain volume) for networks trained using L2, L1, SSIM, 50% L2 & 50% SSIM loss functions. BET indicates training when only extracted brain was used as a ground truth. To ensure reliable metric calculations, the first and last few slices, often characterized by sparse data, were excluded from the analysis.

In the post-hoc ablation comparison, the ACNN and non-attention CNN models showed similar performance for GEPCI-to-FLAIR and GEPCI-to-MPRAGE synthesis. For GEPCI-to-R2*, the ACNN demonstrated substantially lower NMAE for R2* reconstruction compared with the non-attention CNN. The ACNN achieved a NMAE of 0.0127±0.0077, whereas the CNN without attention achieved a NMAE of 0.0368±0.0162. These results suggest improved quantitative accuracy of R2* maps, which rely on subtle echo-dependent variations, when attention mechanisms are incorporated into the network architecture.

### GEPCI to FLAIR and MPRAGE Results

Sixteen test subjects were processed through the GEPCI-to-FLAIR and GEPCI-to-MPRAGE networks. Across these subjects, the AI-generated FLAIR images achieved a mean SSIM of 0.923±0.028, and the AI-generated MPRAGE images achieved a mean SSIM of 0.935±0.022. The individual SSIM values are listed in Table 2. The relatively low SSIM variability across subjects indicates stable network performance. Representative ground-truth and reconstructed images are shown in Fig. 3a-c for FLAIR and Fig. 3d-f for MPRAGE.

**Figure 3.**
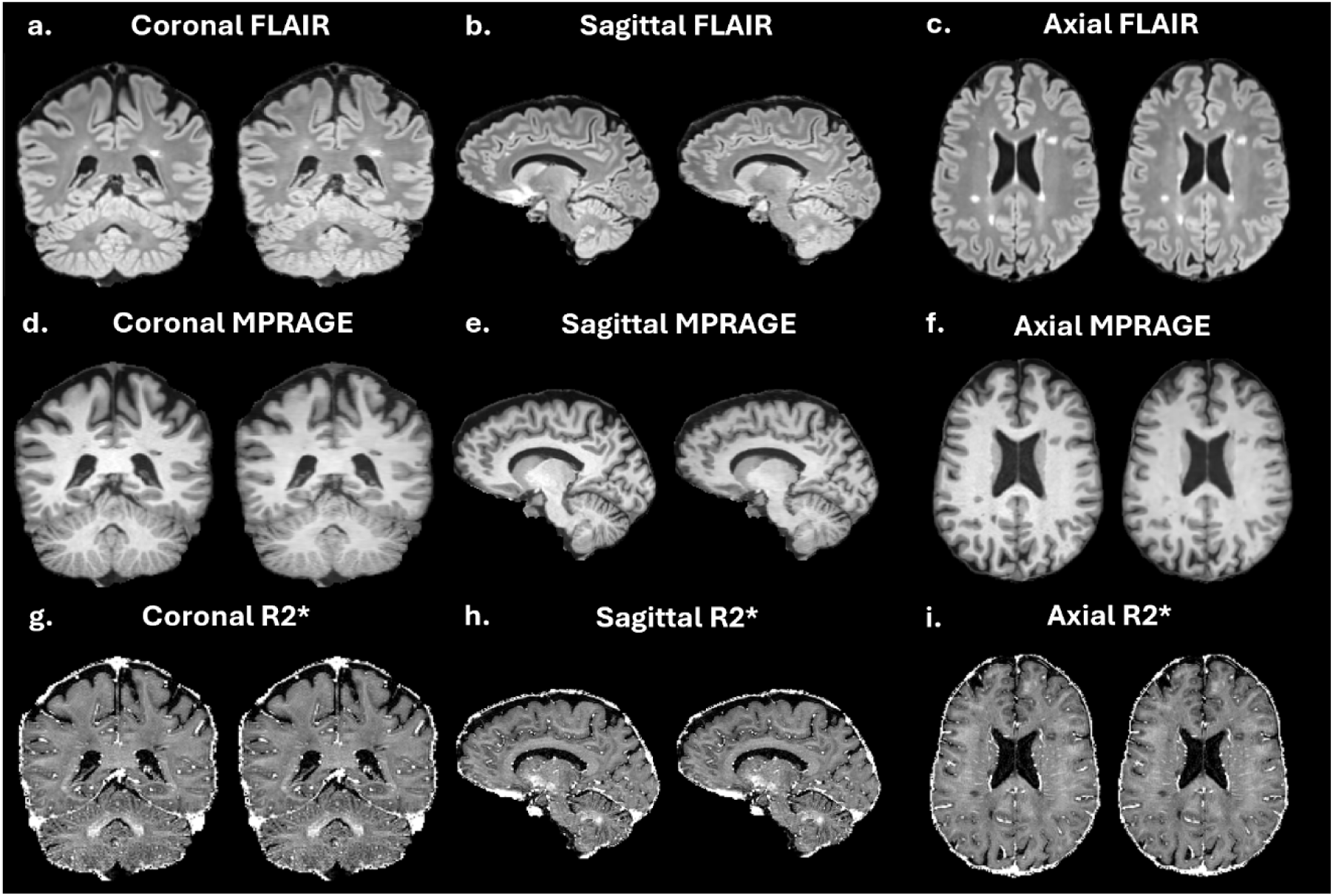
Representative examples of AI-generated GEPCI outputs and corresponding ground-truth images across three anatomical planes. For each contrast, FLAIR (a–c), MPRAGE (d–f), and quantitative R2* maps (g–i), the left image of each pair shows the ground-truth reference and the right image shows the ACNN-generated reconstruction. Across coronal, sagittal, and axial orientations, the AI-generated images closely replicate major anatomical structures and MS-related pathology. Characteristic periventricular and juxtacortical lesions appear as prominent WM hyperintensities in the FLAIR examples and as WM hypointensities in the MPRAGE examples. The AI-generated R2* maps preserve expected tissue contrast patterns and lesion hypointensities consistent with the biophysical model–based reference maps. Together, these examples demonstrate the spatial fidelity of AI-GEPCI for both structural and quantitative imaging and illustrate that lesion morphology and anatomical detail are maintained across orientations.

**Table 2.**
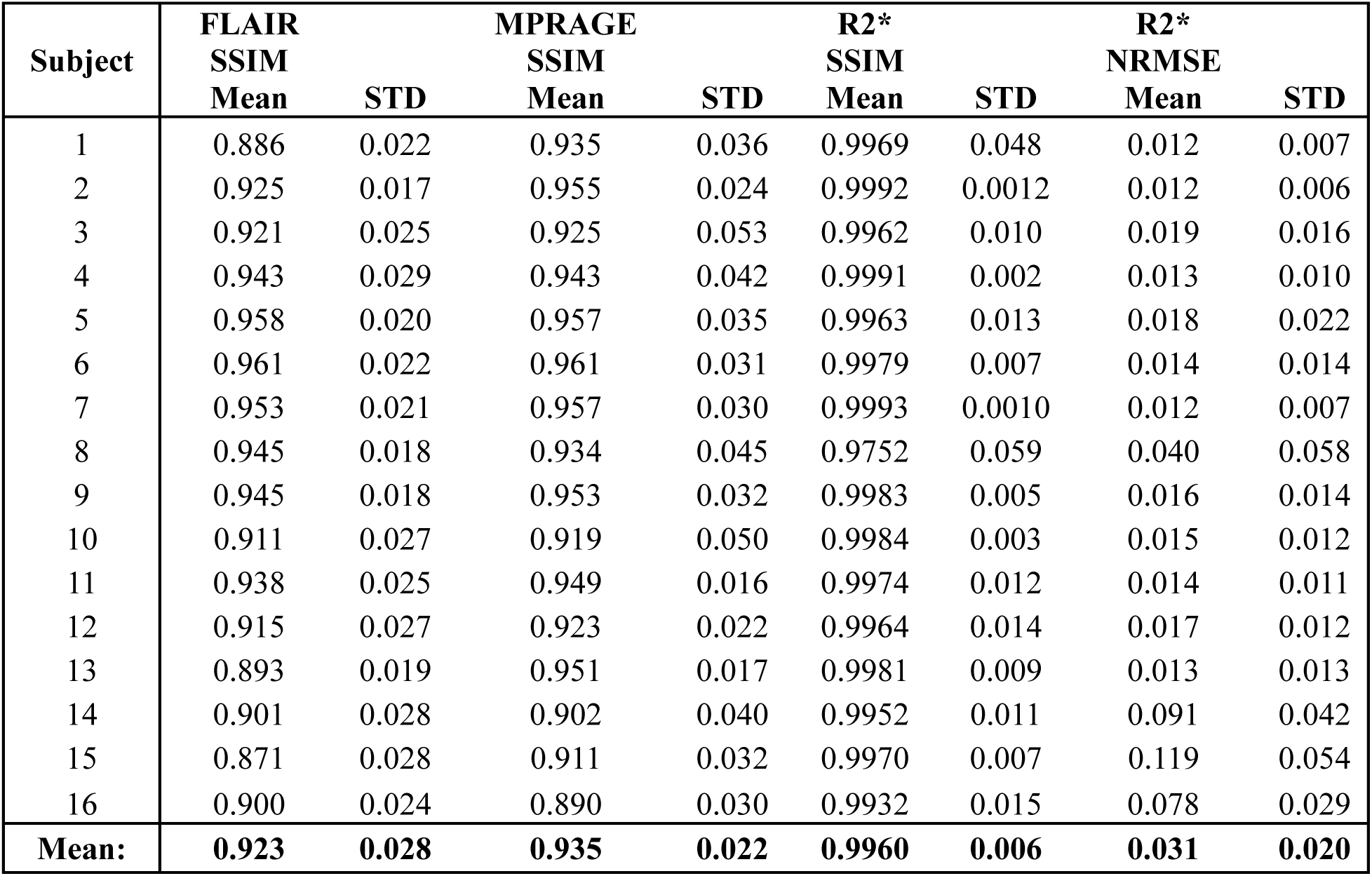
Subject-wise quantitative agreement between AI-GEPCI generated outputs and corresponding native reference images in the hold-out test cohort. For each subject, SSIM is reported as mean ± standard deviation for FLAIR, MPRAGE, and R2* maps; R2* accuracy is additionally summarized by normalized root-mean-square error (NRMSE; mean ± SD). Subjects 1, 10, 13, and 15 had native clinical FLAIR and/or MPRAGE acquisitions with notable artifacts; importantly, AI-GEPCI reconstructions remained clinically interpretable despite these degradations, consistent with the artifact-mitigation behavior.

A small subset of cases exhibited lower SSIM values, corresponding to ground-truth FLAIR images. These, however, were not results of poor ACNN performance, but rather of poor quality of original FLAIR images degraded by motion-related artifacts. Because SSIM directly measures similarity to the available reference images, such artifacts in the ground truth reduce the measured agreement even when the AI-generated reconstructions remain anatomically coherent and clinically interpretable.

### GEPCI to R2* Mapping Results

Across the sixteen test subjects, the AI-generated R2* maps achieved a mean SSIM of 0.996 **±**0.006. Representative ground-truth and reconstructed examples are provided in Fig. 3g–i. As in other contrasts, SSIM was calculated only within the brain, and voxels outside the brain surface were excluded to avoid biasing the similarity measurements.

Quantitative accuracy of the R2* reconstructions was evaluated using NRMSE. The L1-trained network produced a mean NRMSE of 0.031**±**0.02, whereas the L2-trained network yielded a mean NRMSE of 0.055±0.025. These descriptive differences reflect the behavior of the respective loss.

### GEPCI Derived Images

The volumes derived from the G-FLAIR, G-MPRAGE, and G-R2* maps included FLAIR*, MP*, and T2*WI contrasts (Eqs. 2, 3). These derived contrasts were generated across a range of TE values, and representative images for TE = 0.05 sec are presented in Fig. 4 along with corresponding G-FLAIR, G-MPRAGE, and G-R2* maps. Examples corresponding to different TEs are shown in Supplemental Figures 1-3.

**Figure 4.**
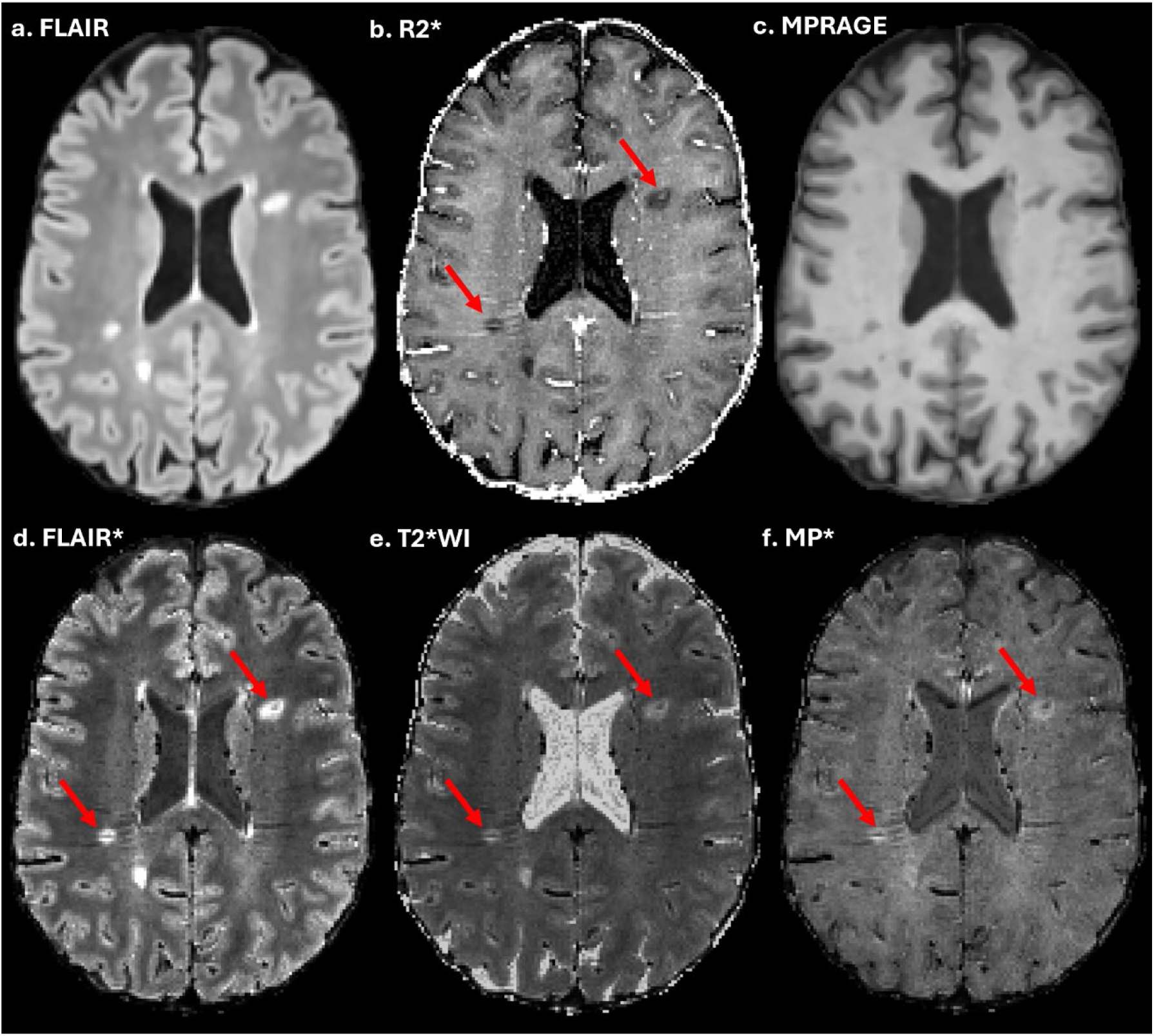
Composite example comprised of the AI-GEPCI generated (a) G-FLAIR, (b) G-R2* map, and (c) MPRAGE images (upper row), and (d) FLAIR*, (e) T2*WI, and (f) MP* images for TE = 50 msec (lower raw). The commonly observed hyperintensity WM periventricular and juxtacortical lesions of MS can be seen in (a), (d), (e), and (f) Similarly, these lesions are seen as hypointensities in (b) and (c) The CVS veins (red arrows) can be seen in the two prominent periventricular lesions in (b), (d), (e), and (f).

T2*WI images highlight regions with lower R2* values, usually corresponding to CSF and lesions. The FLAIR* images suppress the CSF signal, enhance lesions, and provide good contrast between the GM and WM. The MP* images demonstrate enhanced anatomical detail with well-defined lesions and suppressed CSF signal. Note that R2* map, FLAIR*, T2*WI and MP* images may show MS lesions with the CVS or PRL, both are now part of McDonald MS diagnostic criteria (30). CVS appears as a bright structure inside the dark lesion for R2* maps and a dark structure inside the brighter lesion for FLAIR*, MP*, and T2*WI contrasts (Figs. 4, 5). The PRL is a partial or complete iron ring surrounding the lesion borders. The PRL appears as a hypointense ring in the FLAIR*, and T2*WI contrasts (25–31),show a similar appearance on MP*, but manifests as a hyperintense ring in the R2* maps (Fig. 6).

**Figure 5.**
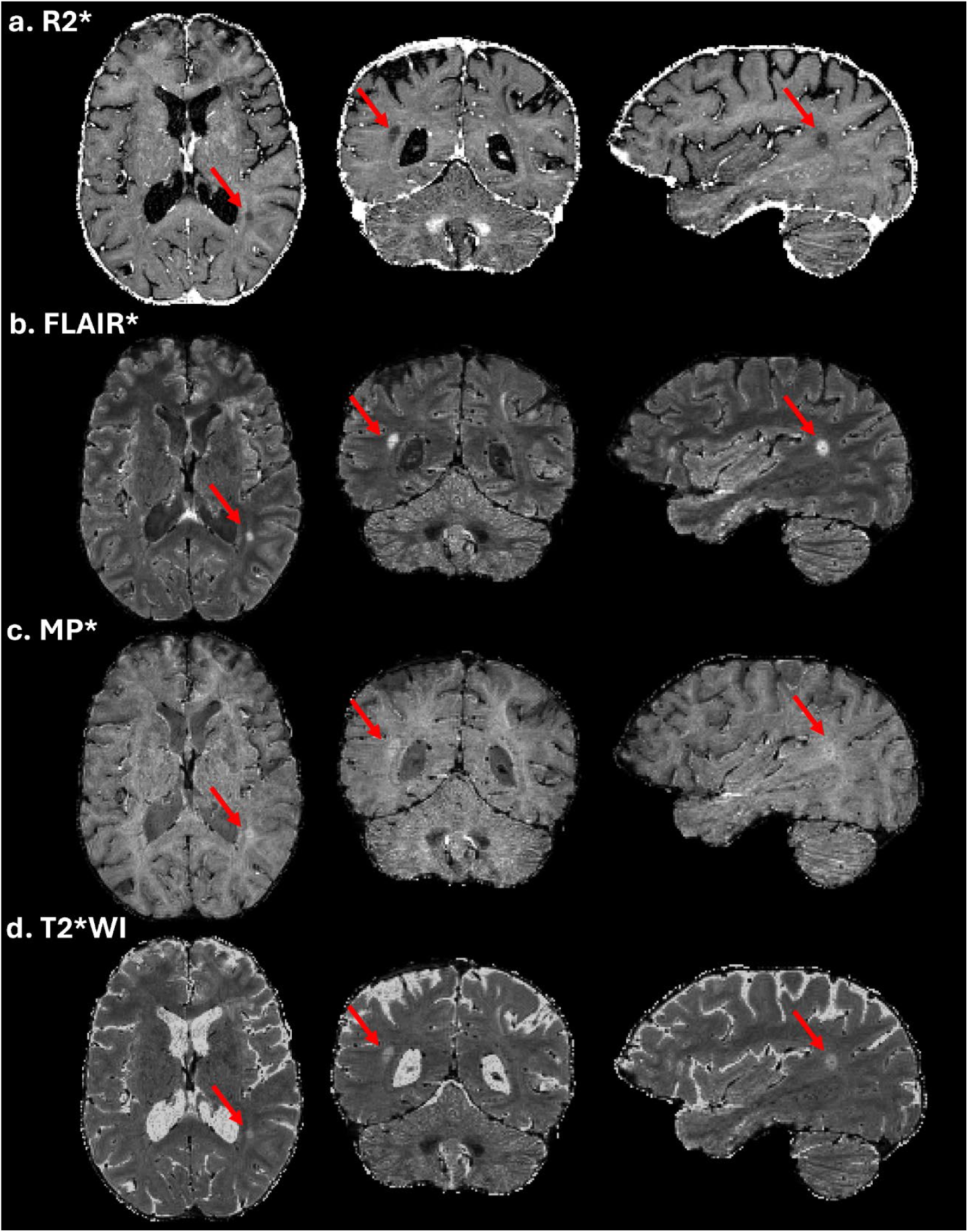
Central vein sign (CVS) example demonstrating the same lesion across axial, coronal, and sagittal planes, visible in all four AI-GEPCI contrasts. (a) G-R2* map, (b) FLAIR*, (c) MP*, and (d) T2*WI images with TE = 50 msec are shown in corresponding views. Red arrows highlight the same CVS lesion in each plane and contrast, confirming its presence and visibility across orientations and modalities. CVS are characteristic of MS pathology (30).

**Figure 6.**
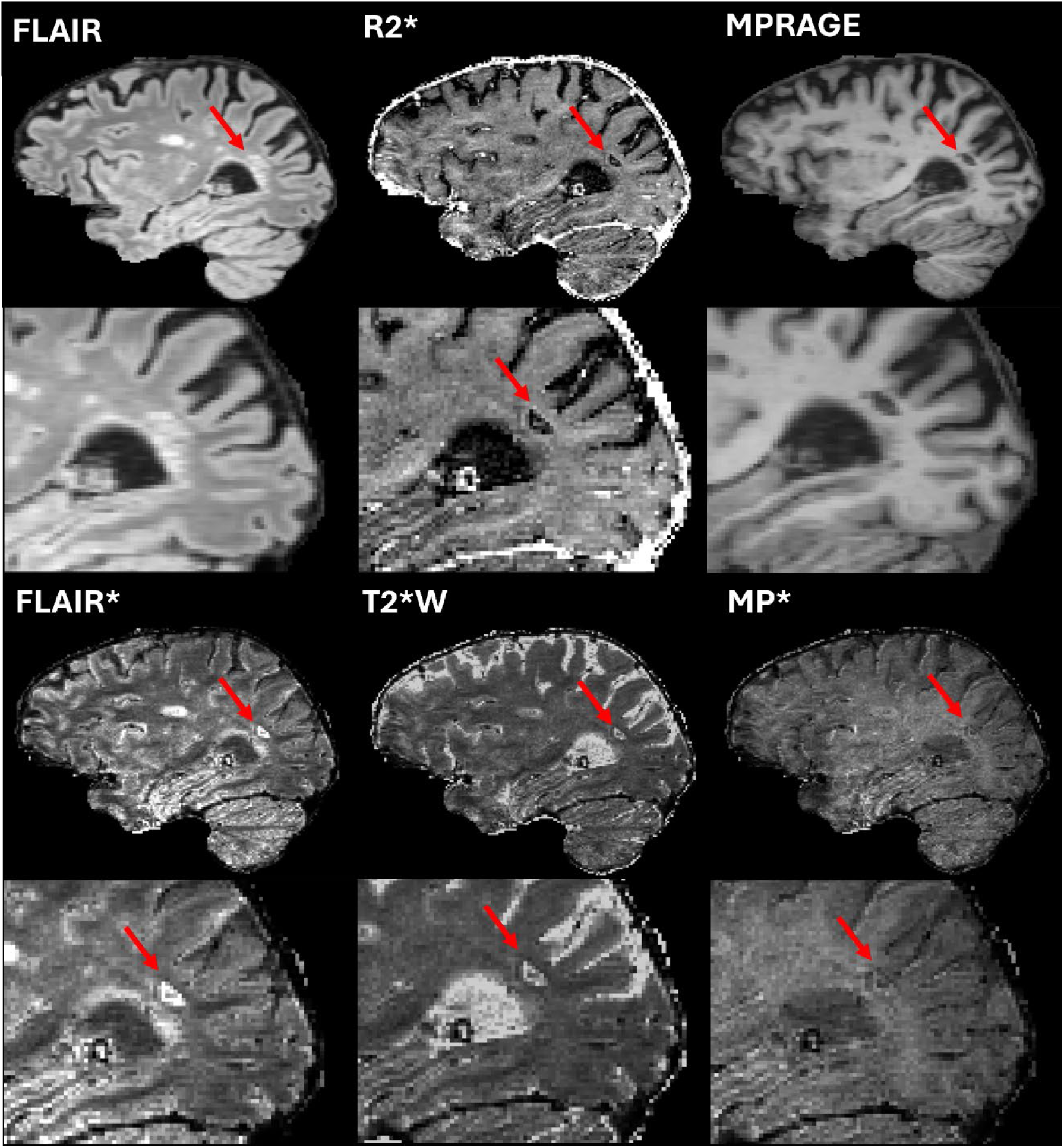
Paramagnetic Rim Lesion (PRL) example demonstrating the same PRL in the sagittal plane across AI-GEPCI generated and derived contrasts. (a) G-FLAIR, (b) R2* map, (c) MPRAGE, (d) FLAIR*, (e) T2*WI, and (f) MP* contrasts, using TE = 50 msec. The PRL is highlighted by red arrows. The PRL border is not visible in (a) FLAIR and (c) MPRAGE. In contrast, (b) shows a thin bright ring around the lesion, while (d), (e), and (f) reveal a dark thin ring around the lesion. PRLs are characteristic of MS, often indicating inflammatory activity and damage to the blood-brain barrier (30).

### Radiological Assessment

The results of expert radiological evaluations are summarized in Table 3. Overall ratings (measured on 1- to-5 scale) were 4.2 for G-FLAIR and 4.5 for G-MPRAGE, both exceeding 4.0 – anchored as the clinically routine imaging quality standard. G-FLAIR and G-MPRAGE ratings were similar for the posterior fossa (4.2 and 4.3, respectively) and artifacts (4.5 for both). Compared to high quality native images used in this study, ratings for the GEPCI AI-generated images were slightly lower (Table 3). The physicians were mixed in identifying which image volume was most likely GEPCI AI-generated. While the vast majority of lesions were equivalently seen on both scans, in some cases small or faint lesion(s) were seen in one scan and not the other. On average, this occurred more often for the ground-truth images than the GEPCI-generated images for FLAIR images (7.3 vs 4.3/16 cases), but rarely for MPRAGE (3.3 vs 1.0/16 cases).

**Table 3.**
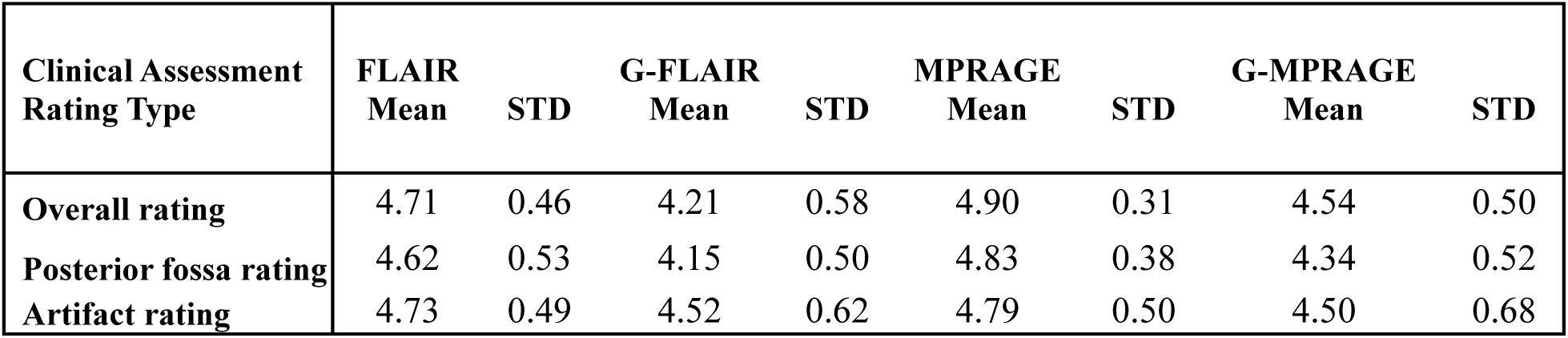
Expert physician assessment of image quality of G-FLAIR, G-MPRAGE and native FLAIR and MPRAGE. Ratings ranged from 1 through 5, with 5 being the best and 4 corresponding to the clinically routine imaging quality standard.

### GEPCI Network Application in MS Lesion Visualization

Following the blinded radiological evaluation, the application of the networks was assessed to identify additional characteristics specific for MS lesions, including CVS and PRL, which are of particularly high clinical utility (30). Periventricular and juxtacortical lesions are a hallmark of MS and are clearly represented in all AI-GEPCI generated and derived contrasts, as shown in Figs. 4 and 5. In addition to these typical MS lesions, CVS lesions are clearly observed across the AI-GEPCI derived results and the G-R2* maps.

According to current criteria, a lesion qualifies as having a CVS if the central vein is visible in at least two imaging planes (30). Similarly, a PRL must be present in at least two imaging planes as well (30). The AI-GECPI results clearly demonstrate lesions that meet these criteria. Fig. 5 presents an example of a CVS lesion visible in all three imaging planes across the G-R2* map and the derived MP*, FLAIR*, and T2*WI images. Fig. 6 illustrates a representative PRL. The PRL appears as a bright partial ring on the G-R2* map (highlighted by a red arrow) and as a dark partial ring on the FLAIR*, MP*, and T2*WI images. These findings demonstrate the network’s ability to replicate clinically relevant imaging biomarkers, including emerging features of high clinical value, supporting its role in MS lesion detection and characterization.

Quantitative comparison of lesion volumes and lesion counts between the AI-GEPCI pipeline and ground truth segmentations demonstrated close correspondence, as illustrated in the scatter plots in Fig. 7. In cases with more complex pathology, such as extensive lesion burden with severe atrophy or a large number of small or closely spaced lesions, automated segmentation was more challenging. In these instances, differences in lesion count primarily reflected lesion confluence or boundary separation rather than missed lesion tissue, resulting in underestimation of lesion number without substantial underestimation of total lesion volume.

**Figure 7.**
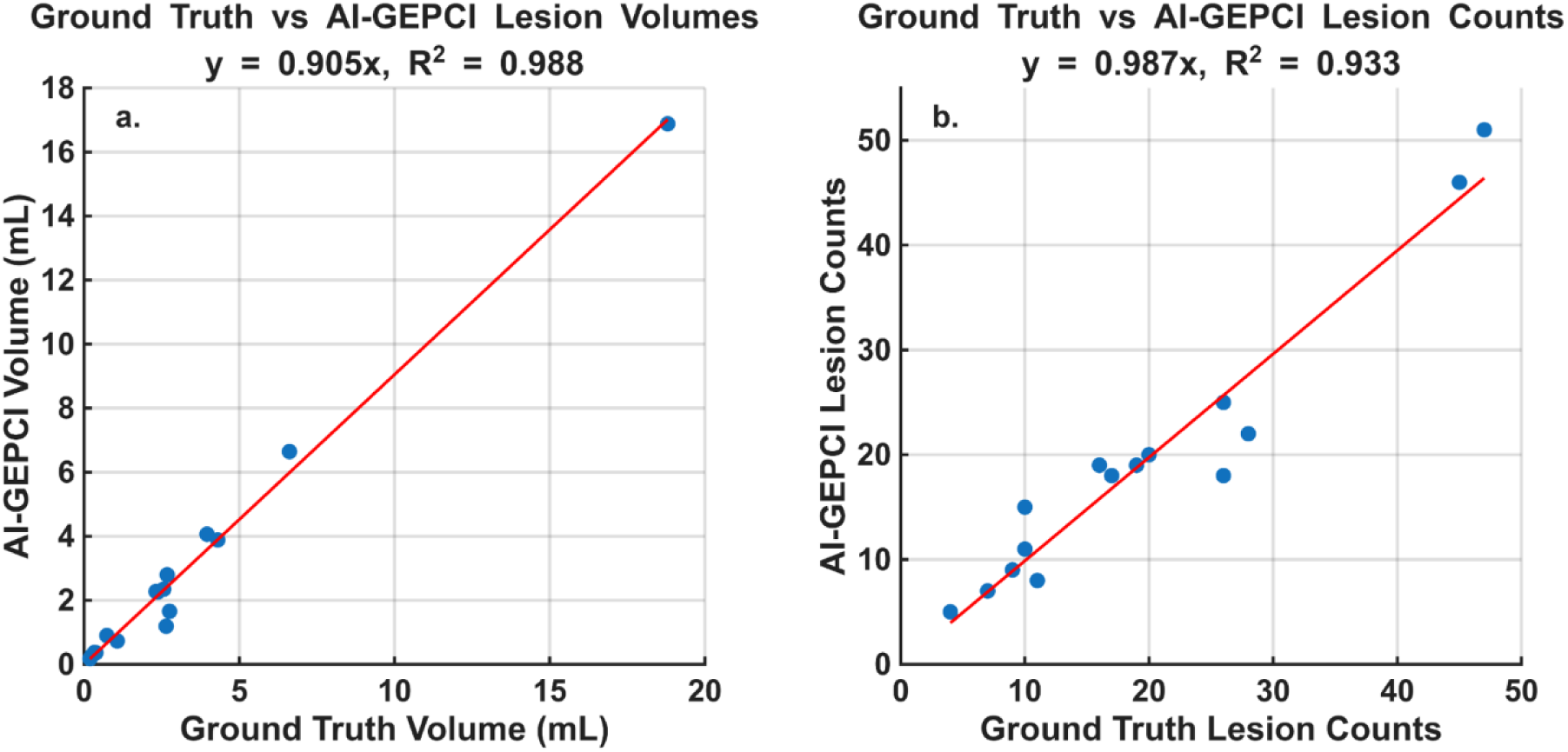
Comparison of AI-GEPCI lesion segmentations to ground truth across lesion volumes and lesion counts. (a) Scatter plot of lesion volume comparison displays the AI-GEPCI (AI) lesion volumes versus the ground truth (GT) lesion volumes. (b) Scatter plot of lesion count comparison from AI and GT segmentations. Each point represents a single test subject. The red lines represent the regression lines. The regressions equations and coefficients of determination (R²) are shown. The high R² values indicate strong correlations.

In addition, a subset of subjects exhibited reduced ground-truth image quality in FLAIR, MPRAGE, or both due to motion or acquisition-related artifacts. These cases coincided with lower SSIM values in Table 2, consistent with reduced agreement between the AI-generated images and degraded reference scans and influenced segmentation-based comparisons. Despite these challenges, lesion volume and lesion count measurements across all subjects showed strong agreement, with coefficients of determination of R² = 0.988 for lesion volume and R² = 0.933 for lesion count.

## Discussion

In this study, ACNNs were developed to generate a suite of naturally co-registered, clinical-quality images from a single mGRE GEPCI acquisition. These include FLAIR, MPRAGE, R2*, T2*WI, FLAIR*, and MP*. The generated images demonstrated excellent agreement with the corresponding clinical images, with SSIM indicating high correspondence for FLAIR, MPRAGE, and R2* maps and also excellent agreement between G-R2* maps and the biophysical model derived reference R2*, (Eq. 1). In blinded assessments, three clinicians judged the GEPCI-derived FLAIR and MPRAGE images to be only slightly lower in quality than the high-quality native FLAIR and MPRAGE images used in this study and found them comparable to or better than clinically routine FLAIR and MPRAGE. Moreover, the GEPCI-derived images performed comparably with regards to detecting lesions while showing utility for CVS and PRL identification. These findings highlight the potential clinical feasibility of this approach. Importantly, this work leverages the inherent efficiency and versatility of GEPCI, reducing acquisition time and ensuring perfect natural co-registration across multiple contrasts, an advantage over existing methods that rely on multiple separate sequences. The ability to generate both structural and quantitative maps from a single acquisition has strong implications for neurological disease imaging, particularly in MS, where accurate lesion visualization, tissue characterization, and quantitative monitoring are critical. Though further optimization of the GEPCI-derived FLAIR and MPRAGE images would be helpful, these results suggest that they might already be clinically suitable. Both L1- and L2-trained networks showed low variability across subjects, although the L1-trained model exhibited lower NRMSE values for R2*, suggesting a possible advantage for this loss function in quantitative mapping.

The ablation analysis supports the use of attention mechanisms for quantitative R2* reconstruction. While both architectures produced visually comparable results, the ACNN achieved markedly lower normalized mean absolute error compared with the non-attention CNN. This suggests that attention gates are particularly beneficial for quantitative GEPCI-derived maps, where accurate modeling of subtle echo-dependent signal variations is critical.

The biophysical foundation of the GEPCI MRI technique lies in its ability to extract a wealth of information from complex mGRE data, which is sensitive to tissue microstructure. By leveraging the variability of contrasts in the mGRE images across multiple gradient echo times, the GEPCI algorithms synthesize these characteristics to generate a plurality of images that highlight different aspects of tissue properties from a single, high-resolution, and fast acquisition. This approach, derived from the mGRE sequence, available on nearly all commercial MRI scanners, could make it widely accessible. The GEPCI method is versatile and produces a suite of naturally co-registered images from a single scan, with low flip angles, thus low specific absorption rates, making it suitable not only for traditional MRI at 1.5T and 3T but also for high-field MRI (7T and above). By retaining complex data, GEPCI can yield additional outputs, included (but not limited to) phase images and QSM maps, as demonstrated in previous work (2, 40). This study focused on GEPCI images derived from mGRE magnitude data, which are widely available on commercial MRI scanners.

The developed ACNNs translate mGRE data into a suite of naturally co-registered GEPCI images, including FLAIR, MPRAGE, R2* map, FLAIR*, MP*, and T2*WI contrasts. This approach addresses the need for accurate and efficiently acquired brain MR images for research and clinical applications. Unlike existing networks relying on multiple scans (e.g., T1, T2, FLAIR) for modality translation, this method requires only a single GEPCI scan, reducing acquisition time and ensuring the intrinsic co-registration of all direct and derived results. By leveraging the advantages of GEPCI combined with deep learning, the AI-powered GEPCI approach offers promising solutions for brain MR imaging. This suite of images is particularly relevant for diseases such as MS, where both gray matter and white matter pathology require sensitive, co-registered imaging for accurate detection and monitoring.

Among the generated contrasts, MP* represents a novel addition that draws conceptual inspiration from FLAIR*, which combines FLAIR and T2*WI imaging to enhance lesion and vein conspicuity in MS. While FLAIR* improves the detection of lesions and the central vein sign through voxel-wise combination of FLAIR and T2*WI data, MP* builds upon structural information from MPRAGE by multiplying it with T2*WI echoes across multiple echo times. This synthesis captures both fine anatomical detail and echo-dependent pathological features, mimicking how FLAIR* highlights lesions and veins, while retaining the spatial resolution and contrast of MPRAGE. Notably, MP* contrast can be modulated by selecting the echo time: using an early echo (e.g., TE ≈ 1 msec) emphasizes MPRAGE-like structural detail, while later echoes (e.g., TE > 30 msec) enhance lesion conspicuity comparable to FLAIR*, offering a flexible diagnostic perspective from the same data. MP* thus offers a complementary and tunable contrast that may enhance visualization of tissue changes relevant to MS. Since MP*, FLAIR*, and T2*WI are all derived from the same single GEPCI acquisition and are inherently co-registered, they collectively provide a rich, multi-contrast framework for lesion characterization. Further studies will be necessary to fully evaluate the diagnostic utility and clinical performance of MP*.

Since 2018, various networks have been developed with the aim of synthesizing various image contrasts. Some models aim at data imputation (41, 42), and direct translations from various contrast combinations (4, 43–48). These approaches typically require multiple scan acquisitions as network inputs. In contrast, MRI methods like MR fingerprinting (1, 49), GEPCI (2), Synthetic MR (50), allow generating images faster, with varying contrasts from a single sequence improving efficiency and consistency of contrast generation. From a clinical perspective, shortening the overall MRI protocol has several benefits, including improving patient comfort, reducing motion artifacts (particularly for later sequences), and potentially allowing time for either repeat or additional MRI sequences to improve the value of the MRI. This is of particular clinical relevance in diseases like MS, where patients may experience discomfort and fatigue, and reducing scan times while maintaining or improving lesion detection capabilities is highly advantageous. This paper focuses on the AI-GEPCI approach and demonstrates that compared to previously published GEPCI image generation approaches (2, 4), the ACNNs developed in this paper enable fast, clinical-quality image production that can be directly implemented on clinical MRI scanners. Unlike traditional multi-sequence protocols, AI-GEPCI enables rapid generation of high-quality structural and quantitative maps, co-registered by design, from a single fast scan, streamlining imaging without sacrificing diagnostic power.

From a practical perspective, the ACNN models developed in this work provide rapid access to clinically relevant MR imaging from a single GEPCI acquisition, which may support early diagnosis, treatment planning, and longitudinal monitoring. In particular, patients stand to benefit from shorter scan durations while retaining efficient brain pathology detection, including CVS, and PRL lesions. Importantly, the natural co-registration of the GEPCI-derived images removes a frequent source of error that is particularly relevant when evaluating CVS or PRL between multiple contrasts. Reduced scan times also can help alleviate patient discomfort and fatigue, which are common concerns, especially in a population that may need frequent imaging. Improved lesion visualization is crucial in MS (19–22), where monitoring disease progression, particularly in terms of new or active lesions, is essential for treatment planning. Additionally, GEPCI-derived images offer the potential for improved tissue characterization and the ability to track microstructural changes over time, offering insights into disease progression that could lead to more personalized therapeutic strategies. As previously demonstrated, the plurality of GEPCI-generated images also makes this approach valuable for a wide range of clinical and research applications beyond MS, including neurodegeneration in Alzheimer’s Disease (9, 10), psychiatric diseases (15), and traumatic brain injury (16). The ability to generate structural and quantitative maps from a single, rapid acquisition may support advancing understanding of these diseases and could inform future approaches to patient care.

### Limitations

This study has several limitations. The dataset was obtained from a single center, using a single vendor, a single scanner, a single field strength and relatively small number of MRI scans. Only the magnitude component of the GEPCI data was used to generate the FLAIR, MPRAGE, R2* maps, T2*WI, FLAIR*, and MP* contrasts; the complex phase data were not explored but will be incorporated in future model development to expand GEPCI-derived outputs. Although the normalization strategy was intended to reduce scanner-specific bias, validation across larger and more diverse datasets will be required to assess broader applicability. Finally, radiological evaluation indicated that the quality of GEPCI-derived FLAIR and MPRAGE images could benefit from further optimization.

### Conclusion

This study demonstrated the successful application of ACNNs to generate a suite of clinically relevant, co-registered MR images from a single GEPCI acquisition. The approach not only reduces acquisition time by eliminating the need to acquire multiple sequences, but also provides a comprehensive set of naturally co-registered imaging contrasts critical for neurological disease diagnosis and monitoring, particularly in conditions like MS. The high agreement with clinical standards and the promising performance in lesion detection suggest that GEPCI-derived images could be a valuable addition to clinical practice, offering a cost-effective and efficient alternative to current multi-scan protocols. Additionally, AI-GEPCI demonstrates sensitivity to detecting CVS and PRLs. While further optimization and validation across larger datasets are necessary, the potential for widespread adoption of this technique is evident. Ultimately, this work paves the way for improved MRI protocols that can enhance patient care by delivering faster, more accurate diagnostic imaging with minimal discomfort. Since mGRE pulse sequences are readily available on practically all commercial MRI scanners, the AI-GEPCI approach has great potential for broad implementation in clinical practice.

## Data Availability

Data generated or analyzed during the study are available from the corresponding author by request

## Abbreviations

GEPCI: gradient echo plural contrast imaging,
qGRE: quantitative gradient recalled echo,
QSM: Quantitative Susceptibility Mapping,
SWI: susceptibility weighted imaging,
ACNN: attention convolutional neural network,
SSIM: structural similarity index measure,
NRMSE: normalized root mean squared error,
NMAE: normalized mean absolute error.

## Data Sharing Statement

Data generated or analyzed during the study are available from the corresponding author by request.

## Supplemental Figures

**Supplemental Figure 1.**
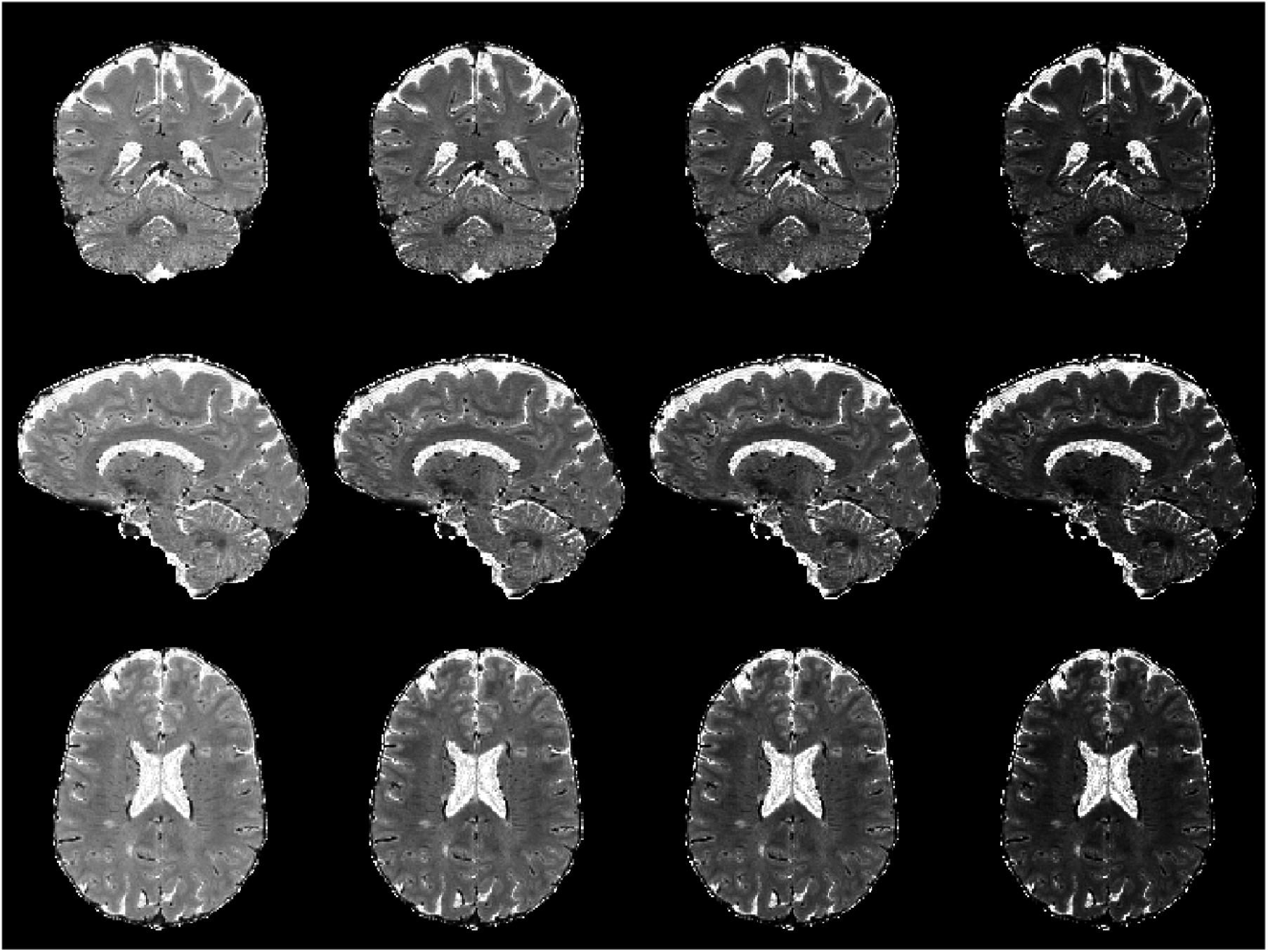
T2*WI example montage. These T2*WI examples are calculated from the generated R2* map results using a TE of 35 msec, 50 msec, 70 msec, and 100 msec for the left column and progressing to the right.

**Supplemental Figure 2.**
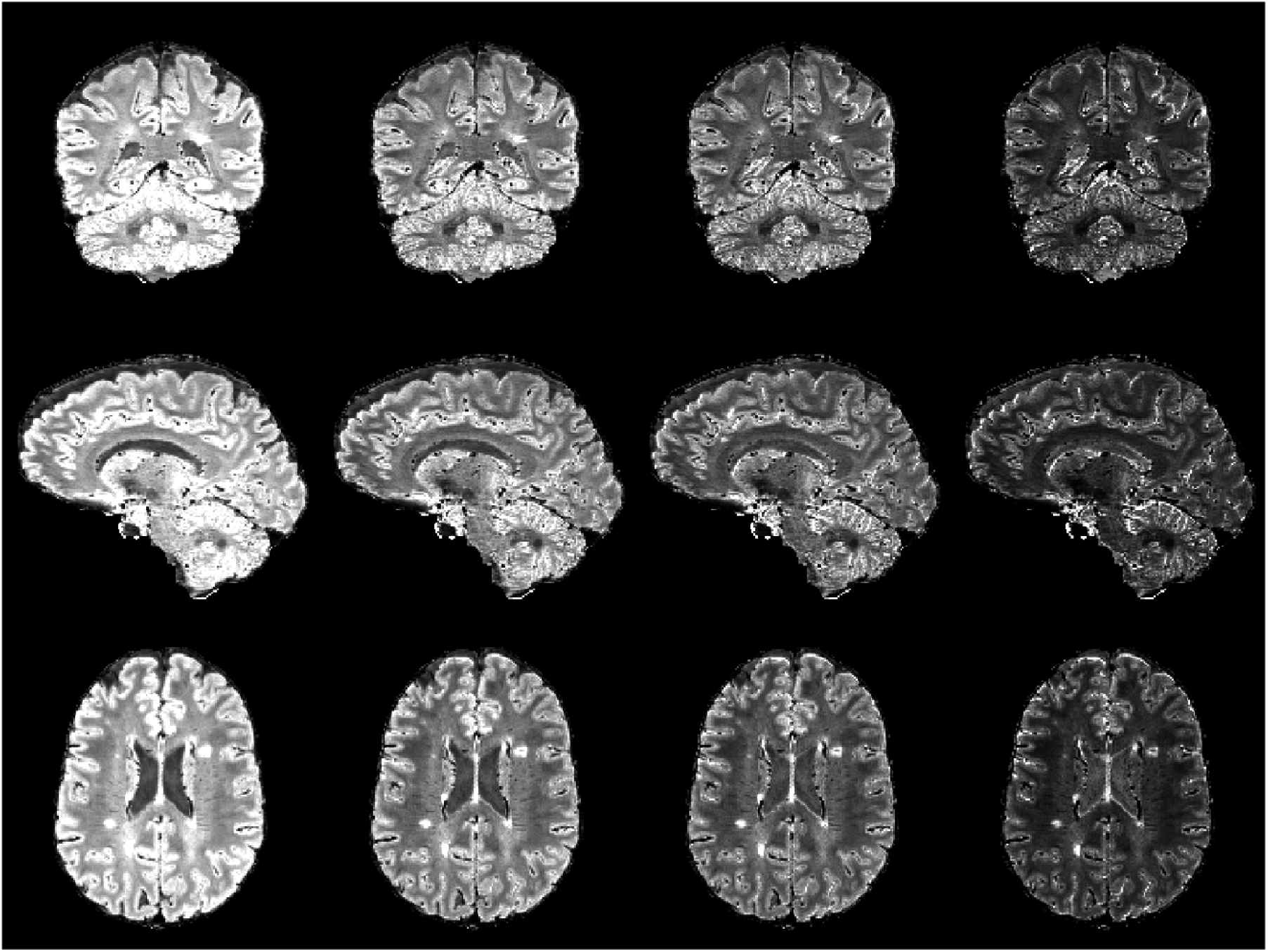
FLAIR* example montage. These FLAIR* examples are calculated from the R2* results using the TE values of 35 msec, 50 msec, 70 msec, and 100 msec. TE increases from the left column to the right column.

**Supplemental Figure 3.**
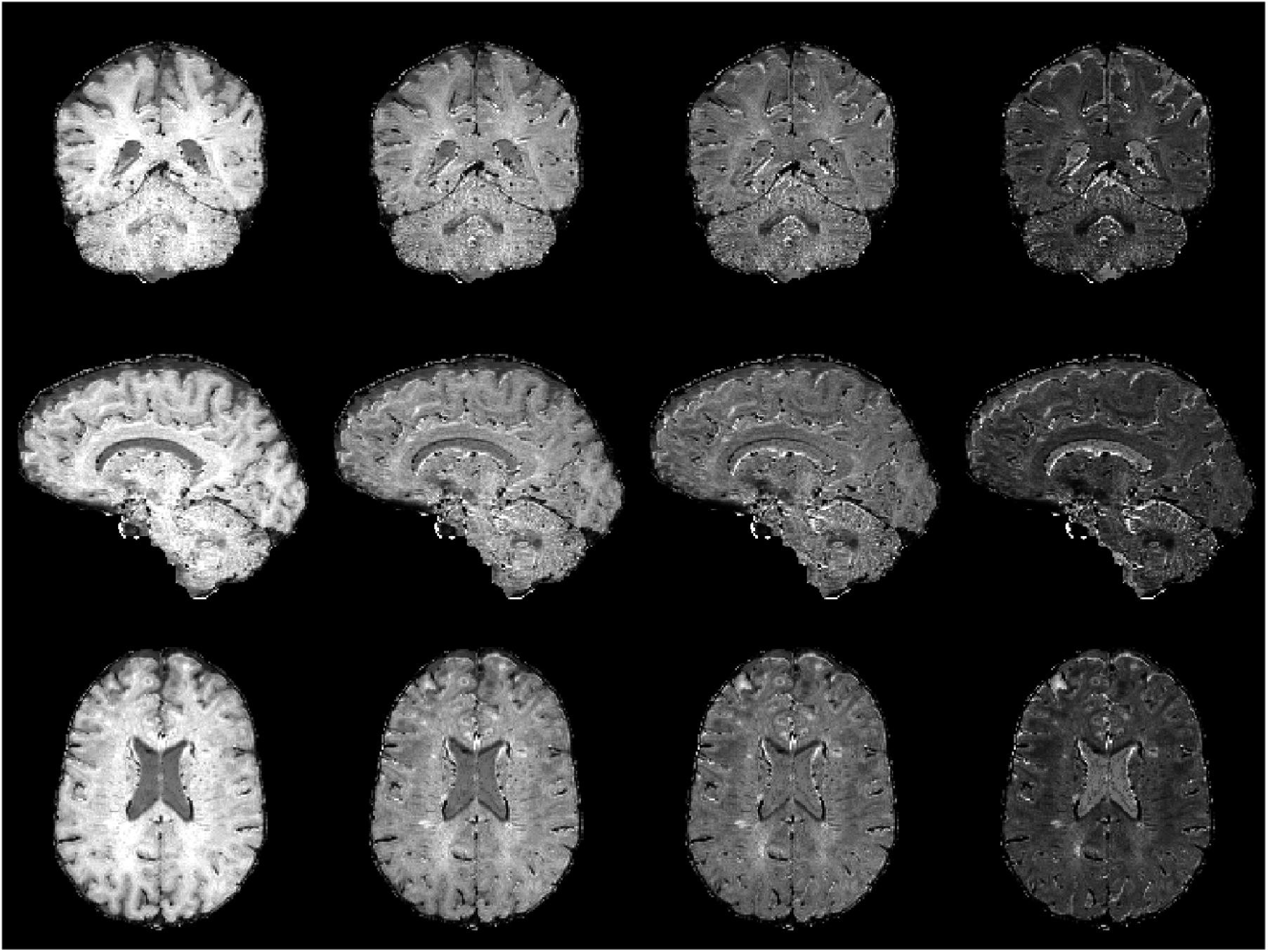
MP* example montage. These MP* examples are calculated from the R2* results using the TE values of 35 msec, 50 msec, 70 msec, and 100 msec. TE increases from the left column to the right column.

